# A panoptic segmentation approach for tumor-infiltrating lymphocyte assessment: development of the MuTILs model and PanopTILs dataset

**DOI:** 10.1101/2022.01.08.22268814

**Authors:** Shangke Liu, Mohamed Amgad, Muhammad A. Rathore, Roberto Salgado, Lee A.D. Cooper

## Abstract

Tumor-Infiltrating Lymphocytes (TILs) have strong prognostic and predictive value in breast cancer, but their visual assessment is subjective. To improve reproducibility, the International Immuno-oncology Working Group recently released recommendations for the computational assessment of TILs that build on visual scoring guidelines. However, existing resources do not adequately address these recommendations due to the lack of annotation datasets that enable joint, panoptic segmentation of tissue regions and cells. Moreover, existing deep-learning methods focus entirely on either tissue segmentation or cell nuclei detection, which complicates the process of TILs assessment by necessitating the use of multiple models and reconciling inconsistent predictions. We introduce *PanopTILs*, a region and cell-level annotation dataset containing 814,886 nuclei from 151 patients, openly accessible at: sites.google.com/view/panoptils. Using PanopTILs we developed *MuTILs*, a neural network optimized for assessing TILs in accordance with clinical recommendations. MuTILs is a concept bottleneck model designed to be interpretable and to encourage sensible predictions at multiple resolutions. Using a rigorous internal-external cross-validation procedure, MuTILs achieves an AUROC of 0.93 for lymphocyte detection and a DICE coefficient of 0.81 for tumor-associated stroma segmentation. Our computational score closely matched visual scores from 2 pathologists (Spearman R=0.58-0.61, p<0.001). Moreover, computational TILs scores had a higher prognostic value than visual scores, independent of TNM stage and patient age. In conclusion, we introduce a comprehensive open data resource and a novel modeling approach for detailed mapping of the breast tumor microenvironment.

## Introduction

Advances in digital imaging of glass slides and machine learning have increased interest in histology as a source of data in cancer studies [1,2]. Tissue morphology contains important prognostic and diagnostic information and reflects underlying molecular and biological processes. This work presents approaches for the computational discovery of interpretable predictive histologic biomarkers, focusing on invasive breast carcinomas and immune response. Histopathology is a medical field where medical experts (i.e., pathologists) examine stained microscopic tissue sections to make diagnostic decisions, most often from tumor biopsies. While much of medicine relies on the clinical examination of patients, histopathology is a visual-focused field, like radiology, where much of the focus is on visual pattern recognition.

The term biomarker refers to a biological feature that we can use to indicate a clinical outcome. For example, prognostic biomarkers are biological features associated with good (or bad) prognosis, while predictive biomarkers predict response to therapy in randomized controlled trials [3]. Typically, when a histologic trait is related to outcomes in cancer, it is incorporated into the grading criteria, though this is not always the case. For example, there has been a strong focus on tumor-infiltrating lymphocytes (TILs) as a prognostic and predictive biomarker in breast cancer and other solid tumors in recent years [4]. This is because TILs infiltration can be a somewhat direct visualization of how well the host (patient) body can respond to the growing tumor by immune cells.

The majority of breast cancers are carcinomas. Based on morphology, breast carcinomas include many variants; the most common are infiltrating ductal carcinoma (which originates from breast duct epithelium) and infiltrating lobular carcinoma (from breast acini/glands) [5,6]. There are numerous morphological elements within a single breast cancer slide. Integrative genomic analysis of breast cancer identified four main subtypes, including Luminal-A, Luminal-B, Her2-Enriched/Her2+, and Basal [7]. These subtypes have distinct alterations and are associated with distinct patient survival prospects [8]. TILs are particularly prognostic and predictive of therapeutic response in basal and Her2+ breast carcinomas [9].

The stromal TILs score is the fraction of stroma within the tumor bed occupied by lymphoplasmacytic infiltrates (**Fig. 1**). TILs are assessed visually by pathologists through examination of formalin-fixed paraffin-embedded, hematoxylin and eosin (FFPE H&E) stained slides from tumor biopsies or resections. They are subject to considerable inter- and intraobserver variability, and hence a set of standardized recommendations was developed by the International Immuno-Oncology Working Group [10,11]. Nevertheless, observer variability remains a critical limiting factor in the widespread clinical adoption of TILs scoring in research and clinical settings. Therefore, a set of recommendations was published for developing computational tools for TILs assessment [12]. A number of existing computational algorithms have been developed to score TILs. However, most diverge from clinical scoring recommendations, as summarized by Amgad *et al.* [12]. This report describes MuTILs, an interpretable deep-learning model for breast cancer WSIs, with a special emphasis on evaluating TILs. MuTILs jointly classifies both tissue regions and individual cell nuclei to produce a panoptic segmentation for TIL scoring and other applications.

**Fig 1.**
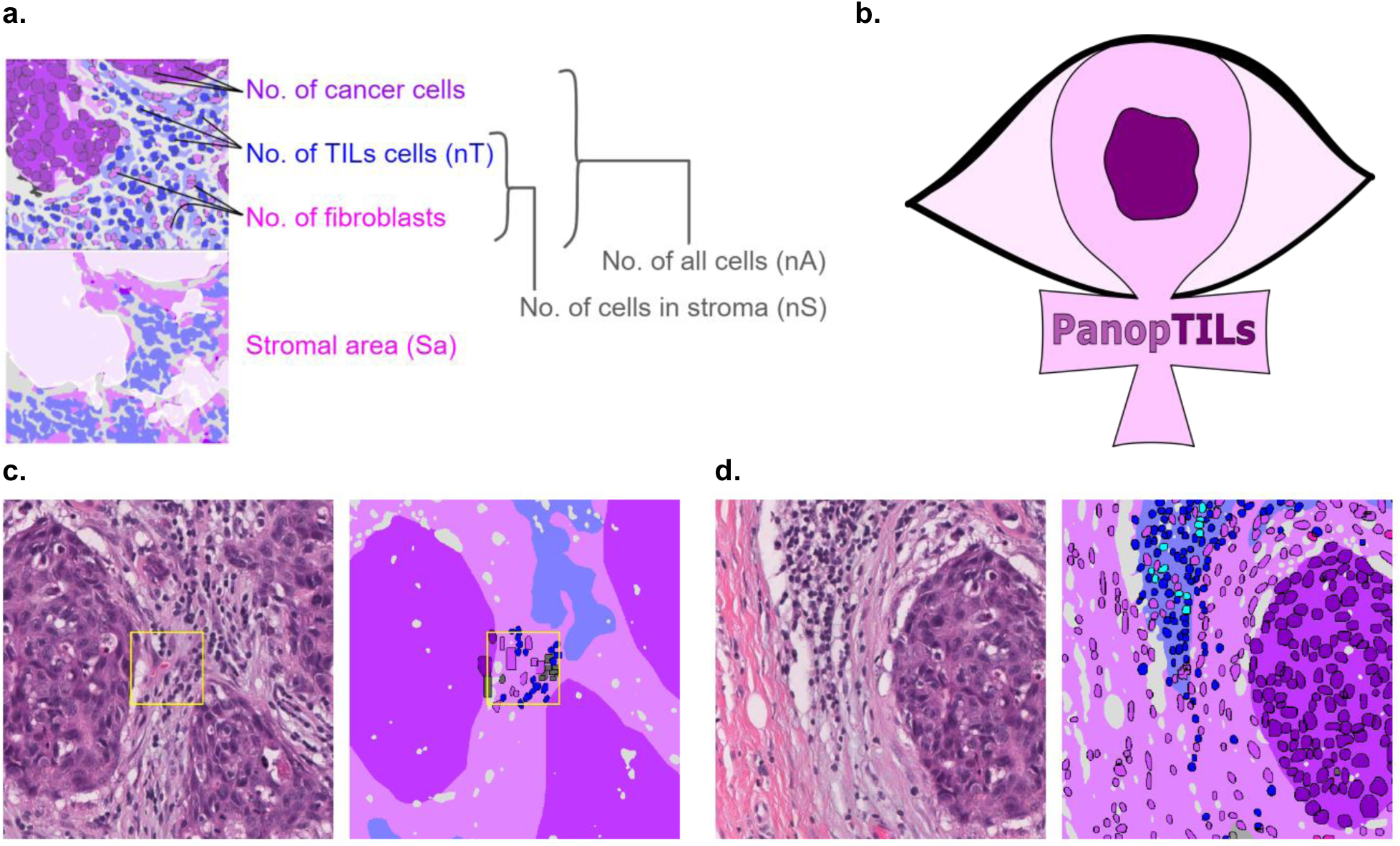
Construction of the PanopTILs dataset to facilitate computational scoring of TILs. **a.** Components of various variants of the computational TILs score. **b.** Logo of our Panoptic segmentation dataset, PanopTILs, which reconciles and expands the region-level and cell-level annotations from the BCSS [17] and NuCLS [18] datasets to better suit the task of densely mapping the tumor microenvironment for TILs assessment. PanopTILs is openly accessible at: sites.google.com/view/panoptils. **c.** The result of combining manual tissue and nucleus annotations from the BCSS and NuCLS datasets. This variant of PanopTILs was used for calculating validation accuracy metrics for our panoptic segmentation model. **d.** Expansion of the manual nuclei annotations to facilitate panoptic (MuTILs) model training. This expansion was done by training additional models to extrapolate nuclei annotations beyond the manual annotations as in [19]. These extrapolated data were used in MuTILs model training and were not used in validation.

## Methods

MuTILs jointly classifies tissue regions and cell nuclei and extends our earlier work on this topic (**Fig. 2**) [13]. It acts as a panoptic segmentation algorithm; that is, it uses semantic segmentation to delineate tissue regions and instance segmentation to segment and classify individual cell nuclei to enable a holistic, context-aware assessment of TILs [14]. MuTILs comprises two parallel U-Nets (each with a depth of 5) for segmenting tissue regions and nuclei at 10X objective and 20X objective magnifications, respectively [15]. Inspired by the HookNet method, information is shared from the tissue region segmentation to inform nucleus segmentation by providing low-power context [16]. Additionally, region predictions from the low-resolution branch are upsampled to 20X magnification and used to constrain the predicted nucleus classes. Tissue region predictions are used to infer attention maps that define the likelihood of different nuclei types occurring based on learned prior probabilities. These attention maps also incorporate user-defined compatibility kernels that prohibit biologically implausible predictions, for example, a fibroblast nucleus in a tumor tissue region. The MuTILs model was trained using a multi-task loss that gives equal weight to Regions of Interest (ROI) ROI and High-Power Field (HPF) region predictions, unconstrained HPF nuclear predictions, and region-constrained nuclear predictions.

**Fig 2.**
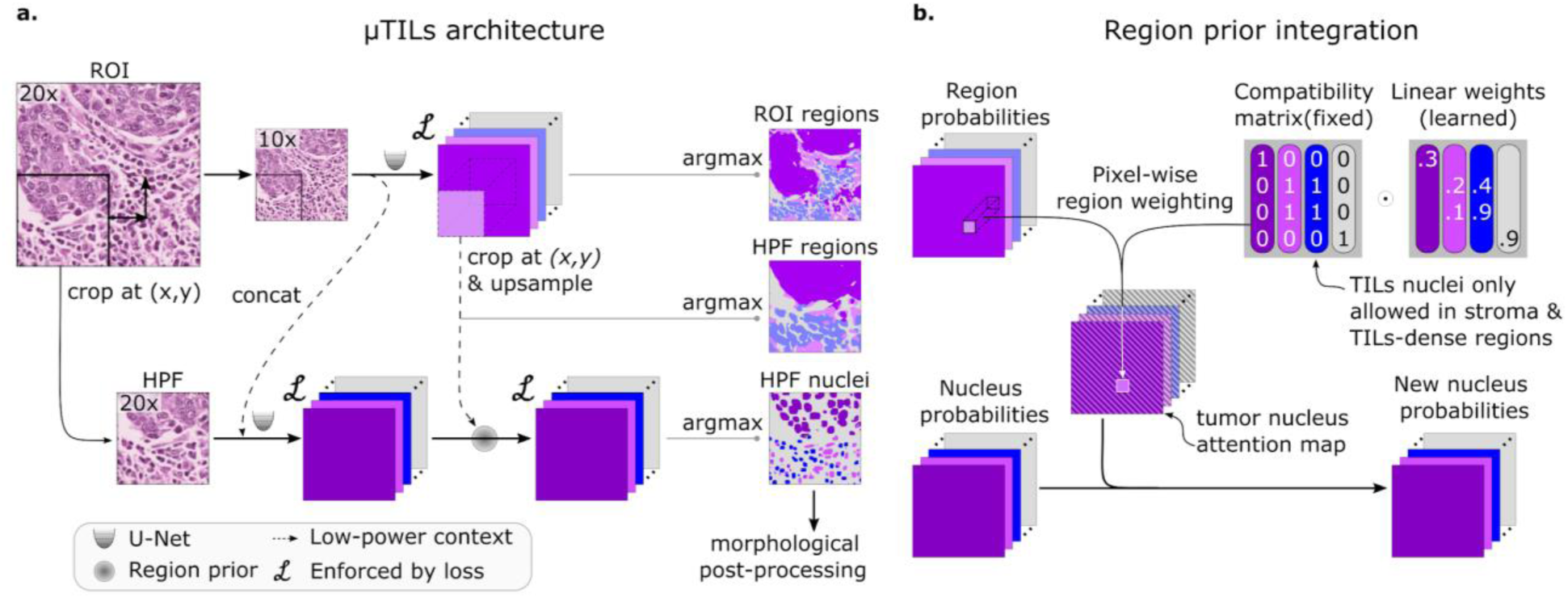
MuTILs model architecture. **a.** The MuTILs architecture utilizes two parallel U-Net models to segment regions at 10X objective magnification and nuclei at 20X objective magnification. Inspired by HookNet, we passed information from the low-resolution region segmentation branch to the high-resolution nuclei classification branch by concatenation. This concatenation, as indicated by the dashed arrow, enriches the high-resolution data with contextual details. Additionally, region predictions from the low-resolution branch are upsampled and used to constrain the possible nucleus classifications in the high-resolution branch. The model was trained using a multi-task loss that gives equal weight to ROI and HPF region predictions, unconstrained HPF nuclear predictions, and region-constrained nuclear predictions. **b.** Region predictions are used to constrain nucleus predictions to enforce compatible cell type predictions through class-specific attention maps. These maps represent the likelihood for each nuclei class occurring at different points in space based on the region prediction, user-defined hard constraints on what cell types can occupy what tissue regions, and learned prior probabilities describing cell type and region type associations. Hard constraints can be used to define rules that prohibit, for example, a nucleus from being classified as a fibroblast within a tumor region.

We created a panoptic segmentation dataset that fuses the annotations from two public datasets: the Breast Cancer Semantic Segmentation dataset (BCSS) [17] and the Nucleus classification, localization, and segmentation dataset (NuCLS) [18]. These datasets were produced through a crowdsourcing process that engaged an international network of medical students, pathology residents, and pathologists using a web-based platform as described in [17, 18]. These datasets annotated regions selected in WSIs from 125 infiltrating ductal breast carcinoma patients from The Cancer Genome Atlas. We call this combined dataset *PanopTILs*, since it enables the panoptic segmentation of tissue regions and cell nuclei necessary for TIL assessment (**Fig 1**). The PanopTILs dataset contains manual annotations comprising 16,322 cancer cells, 9,596 lymphocytes, 6,945 fibroblasts, 5,943 debris, and 4,641 plasma cell nuclei (see Table S1), along with semantic tissue annotations within 1,317 regions of interest where cancer and normal epithelium, stroma, immune infiltrates, and necrosis were annotated. Manually annotated nuclei are concentrated in 256×256 pixel regions-of-interest (0.5 MPP resolution, 20X objective) centered within a larger 1024×1024 pixel area defining the semantic segmentation (see **Fig. 3**, center).

**Fig 3.**
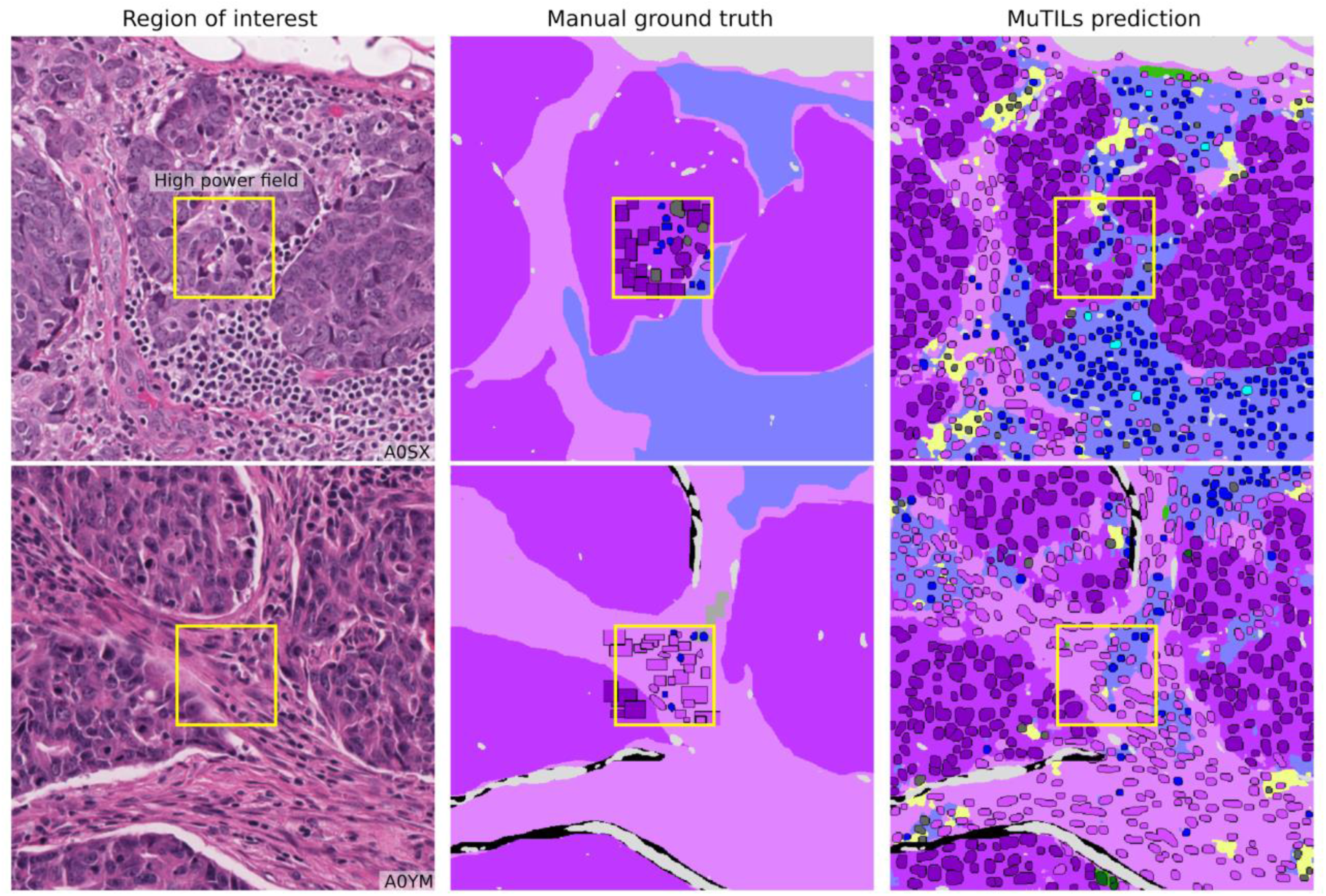
Reconciliation of manual region and nucleus ground truth to produce the PanopTILs validation dataset. Each high-power field from the pathologist-corrected single-rater NuCLS dataset was padded to 1024×1024 at 0.5 MPP resolution (20x objective). As a result, each ROI had region segmentation for the entire field (from the BCSS dataset) and nucleus segmentation and classification for the central portion (from the NuCLS dataset). Note that the nucleus ground truth contains a mixture of bounding boxes and segmentation. The fields shown here are from the testing sets.

**Table 1.** Generalization accuracy for region segmentation and nucleus classification using manual ground truth. Results are on testing sets from the internal-external 5-fold cross-validation scheme (separation by hospital). Fold 1 contributed to hyperparameter tuning, so it is not included in the mean and standard deviation calculation. MuTILs achieves a high classification performance for components of the computational TILs score. Region segmentation performance is variable and class-dependent, with the predominant classes (cancer, stroma, and empty) being the most accurate. The region constraint improves nuclear classification AUROC by ~2-3% overall, mainly by reducing the misclassification of immature fibroblasts and large TILs/plasma cells as cancer (see qualitative examination Figure 7). Classes and the simplified merged classes are indicated in the first and second columns respectively. * Classes that contribute to the computational TILs score. † Performance for Necrosis/Debris and TILs-dense regions is modest, primarily because of the inherent subjectivity of the task and variability in the ground truth. Infiltrated stromal regions do not have clear boundaries and necrotic regions also often have TILs infiltrates at the margin or adjacent areas of fibrosis, which are inconsistently labeled as necrosis, stroma, or TILs-dense in the ground truth. Nonetheless, classifying cells/material that comprise necrotic regions (neutrophils, apoptotic bodies, debris, etc.) is reasonable at higher magnification. ‡ The model fails to distinguish normal and neoplastic breast epithelium at 10x magnification. This failure is likely caused by: 1. The low representation of normal breast tissue in the validation data from NuCLS and BCSS datasets; 2. Inconsistency in defining “normal,” which is sometimes used in the sense of “non-cancer” (including benign proliferation), and sometimes only refers to terminal ductal and lobular units (TDLUs). At high resolution, the distinction between cancer versus normal/benign epithelial nuclei is reasonable.

| Class | Merged class | Fold 1 | Fold 2 | Fold 3 | Fold 4 | Fold 5 | Mean | Std |
| --- | --- | --- | --- | --- | --- | --- | --- | --- |
| **Regions at 10x objective (DICE)** |  |  |  |  |  |  |  |  |
| Cancer | Epithelium | 84.4 | 82.1 | 83 | 82.8 | 82.8 | 82.7 | 0.4 |
| Normal ‡ | Epithelium | 1.6 | 2.3 | 2.1 | 2.3 | 2.3 | 2.3 | 0.1 |
| Stroma * | Stroma | 81.3 | 80.2 | 81 | 80.8 | 81 | 80.8 | 0.4 |
| TILs † | TILs | 64.8 | 64 | 65.3 | 65.6 | 65.6 | 65.1 | 0.8 |
| Necrosis/Debris † | Necrosis/Debris | 64.1 | 55.6 | 56.7 | 57.3 | 57.1 | 56.7 | 0.8 |
| Empty | - | 83.5 | 83.5 | 84 | 84.2 | 84.3 | 84.0 | 0.4 |
| **Nuclei at 20x objective (AUROC)** |  |  |  |  |  |  |  |  |
| Cancer | Epithelial | 96.5 | 97.2 | 98 | 97.4 | 91.1 | 95.9 | 3.2 |
| Normal ‡ | Epithelial | - | 84.6 | 89.3 | 80 | 74.7 | 82.2 | 6.3 |
| Fibroblast * | Stromal | 90.4 | 93 | 91.8 | 93.5 | 85.8 | 91.0 | 3.6 |
| Lymphocyte * | TIL | 93.3 | 92.3 | 93.6 | 91.9 | 94.2 | 93.0 | 1.1 |
| Plasma Cell * | TIL | 80.9 | 73.5 | 88 | 78.9 | 85.8 | 81.6 | 6.6 |
| Debris † | Debris | 82.8 | 84.9 | 80.1 | 93.9 | 57.1 | 79.0 | 15.7 |
| Micro-avg. | - | 91.9 | 92.2 | 95.6 | 93.5 | 88.9 | 92.6 | 2.8 |
| Macro-avg. | - | 85.4 | 83.9 | 86.3 | 85.2 | 75.3 | 82.7 | 5.0 |
| **Nuclei without region constraint (AUROC)** |  |  |  |  |  |  |  |  |
| Micro-avg. | - | 90.5 | 91.1 | 95.4 | 91.9 | 86.2 | 91.2 | 3.8 |
| Macro-avg. | - | 84.5 | 78.1 | 86.9 | 81.5 | 73.1 | 79.9 | 5.8 |

For the purposes of training MuTILs models, nuclei annotations were extrapolated to the full 1024×1024 ROI using models for nuclear instance classification from [19] to infer nuclear boundaries and classes in the periphery. The extrapolation models were trained using the manual nuclear annotations from the central 256×256 regions of training images and then applied to margins of the 1024×1024 ROI to infer nuclei annotations there (see Figure 1d). During MuTILs model training, we also supplement PanopTILs with annotations from 85 slides from the Cancer Prevention Study II cohort in order to enrich the training data with lower-grade and normal tissue examples (these annotations are not included in the PanopTILs release) [20]. Slides were separated into training and testing sets using 5-fold internal-external cross-validation, using the same folds described in [18,21]. Model performance was assessed entirely on manual cell annotations (**Fig. 3**). Extrapolated nuclei annotations were not used for validation. The fields depicted in **Fig. 3** are from the application testing sets.

The classes in the PanopTILs training set are more granular than what is required for TIL scoring (for example, discriminating lymphocyte from plasma cell nuclei). Some of these finer subclassifications have lower inter-rater agreement in the annotation datasets (“unreliable”), or are not abundant enough for a model to learn (normal epithelium). Therefore, we assessed performance by grouping several classes to form a more reliable and practical ground truth (epithelium, stroma, TILs). Predictions for normal and cancer epithelium are combined into a single “epithelium” class.

For whole-slide image (WSI) inference, we relied on data from 305 breast carcinoma patients for validation, 269 of whom were infiltrating ductal carcinomas, and 156 were Her2+. Visual scores were assessed by two pathologists and used as the baseline. Scores were performed in accordance with recommendations of the International TILs Working Group, which recommends scoring stromal TILs as a percentage of the stromal areas between nests of carcinoma cells [10]. Scoring was performed within the border of the invasive cancer, but areas occupied by malignant cells are not included in the total assessed area. All mononuclear cells are scored (including both plasma cells and lymphocytes). Pathologists were blinded to each others’ scores, and the scores of the algorithms in these experiments. Scores from reader 1 were used in clinical correlations as this reader is a breast cancer subspecialist.

The WSI accession and tiling workflow used the *histolab* and *large_image* packages and included: 1. Tissue detection; 2. Detection and exclusion of empty space and markers/inking; 3. Tiling the slide and scoring tiles at a very low resolution (2 MPP); 4. Analyzing the top 300 tiles [22,23]. Fixing the number of analyzed ROIs ensured a near-constant run time of less than two hours per slide. Low-resolution tiles with a high composition of cellular (hematoxylin-rich) and acellular (eosin-rich) regions received a higher informativeness score. This favored tiles with more peritumoral stroma. Color deconvolution was performed using the Macenko method from the HistomicsTK package [24,25]. Each of the top informative tiles was assigned one of the trained MuTILs models in a grid-like fashion. This scheme acted as a form of ensembling without increasing the overall inference time.

Trained MuTILs models were then used to segment tissue and nuclear components. A Euclidean distance transform was applied to detect stroma within 32 microns from the tumor boundary. The fraction of image pixels occupied by this peritumoral stroma was used as a saliency score. We assessed the following variants of the TILs score (**Fig. 1**):

1. Number of TILs / Stromal area (nTSa)
2. Number of TILs / Number of cells in stroma (nTnS)
3. Number of TILs / Number of cells anywhere (nTnA)

We obtained these score variants using two aggregation strategies: 1. Globally (aggregating region and nuclear counts from ROIs) and 2. By saliency-weighted averaging ROIs. A simple linear calibration was used to map the computational scores to a similar range as the visual scores to aid interpretation. This calibration first z-scores the visual and computational scores to identify outliers where disagreement is greater than 1.96 standard deviations. The remaining inliers are used to calibrate the computational scores to the visual scores using linear regression with no intercept.

Validation of MuTILS was performed using a strict internal-external 5-fold cross validation [21]. This ensures that slides from a single hospital never appear in both training and testing to better estimate generalizability. In all experiments fold 1 contributed to hyperparameter tuning and so it is not included in reporting of mean and standard deviation for performance metrics. Metrics calculated include the Sørensen–Dice (DICE) coefficient for tissue segmentation, and accuracy, area under ROC curve (AUROC), sensitivity and specificity, precision and recall, F1 score, and Matthews correlation coefficient (MCC) for nucleus classification.

Clinical data analysis used progression-free interval (PFI) as the endpoint used per recommendations from Liu *et al.* for TCGA, with progression events including local and distant spread, recurrence, or death [26]. Kapan-Meier curves were examined for patient subgroups using a TILs-score threshold of 10% for stromal TILs scores. While different thresholds are used in the literature, a 10% is often the defining threshold for a low TIL-score. For the nTnA score variant, a threshold of 3% is used to adjust for the larger number of cells included in the denominator (see Supplementary Figure S1). To avoid having all conclusions rely on a specific choice of threshold, TIL-scores were also included in Cox regression analysis as continuous variables.

## Results

MuTILs has a strong emphasis on explainability; it segments individual regions and nuclei, which are then used to calculate the computational scores. **Table 1** shows the region segmentation and nucleus classification accuracy on the testing sets. MuTILs achieves a high classification performance for components of the computational TILs score, including stromal region segmentation (DICE=80.8±0.4) as well as the classification of fibroblasts (AUROC=91.0±3.6), lymphocytes (AUROC=93.0±1.1), and plasma cells (AUROC=81.6±6.6). Region segmentation performance is variable and class-dependent, with the predominant classes (cancer, stroma, and empty) being the most accurate. The region constraint improves nuclear classification AUROC by ~2-3% overall, mainly by reducing the misclassification of immature fibroblasts and large TILs/plasma cells as cancer. A detailed performance analysis of the impact of region constraint is presented in Supplementary Materials (Supplementary Figure S2 and Tables S2–S7). The generalization accuracy of MuTILs predictions is also supported by a qualitative examination of model predictions on the ROIs from BCSS and NuCLS datasets (**Fig. 3**) and the full WSI (**Fig. 4**). Note that in Fig. 4, the predictions show full WSI inference for illustration.

**Fig 4.**
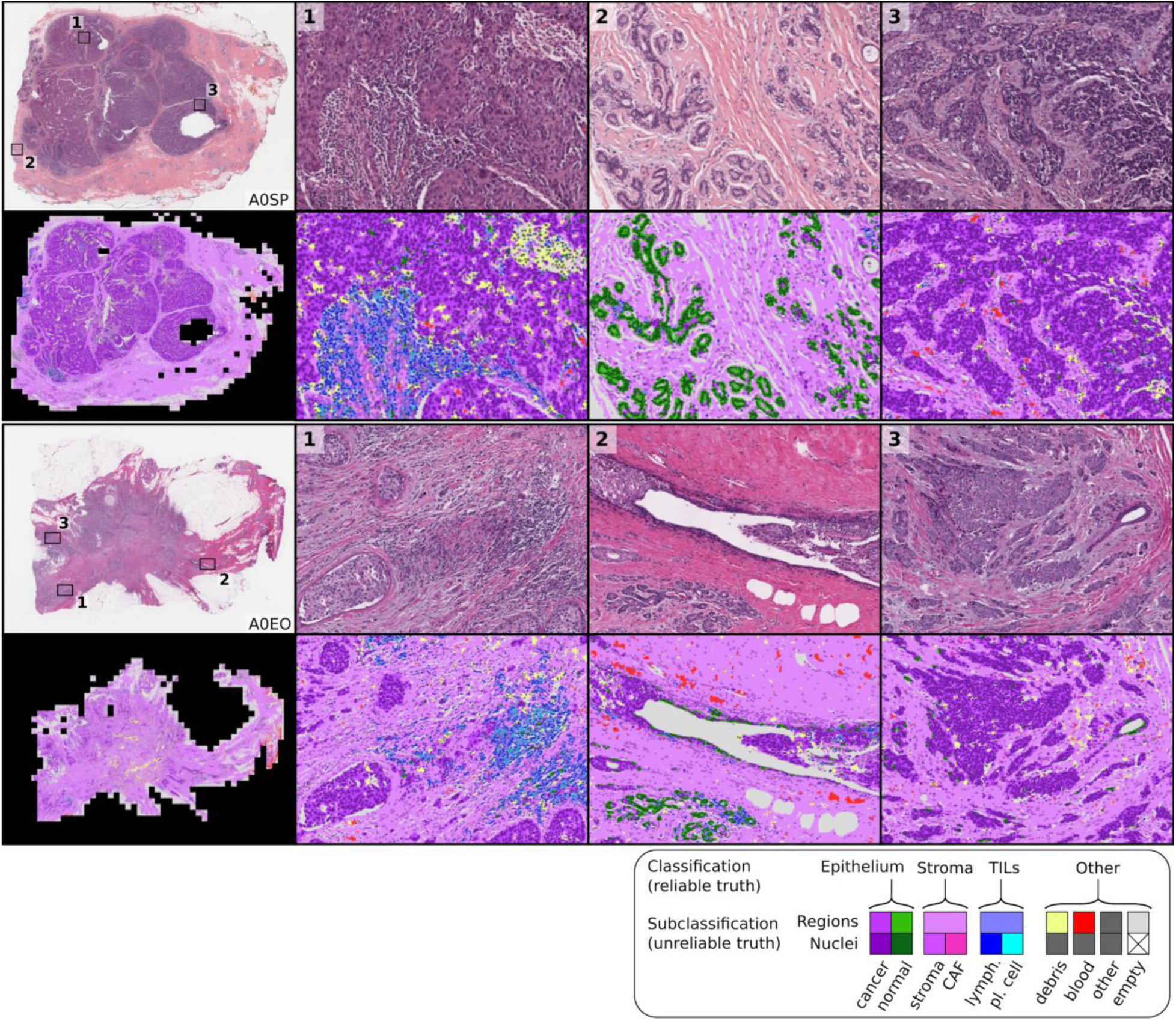
Sample whole-slide predictions from trained MuTILs models. The predictions show full WSI inference for illustration, however, our analysis only admitted the 300 most informative ROIs to the MuTILs model to limit run time to two hours per slide for practical applicability. ROI saliency was measured at a very low resolution (2 MPP) during WSI tiling and favored ROIs with more peritumoral stroma. The training set annotations of cells and tissue regions are more granular than what is necessary for TIL scoring, for example distinguishing between lymphocytes and plasma cells. Some granular classifications have lower inter-rater agreement (“unreliable”), or are not abundant enough for a model to learn. Therefore, we assessed performance by grouping several classes to form a more reliable and practical ground truth (epithelium, stroma, TILs).

**Table 2.**
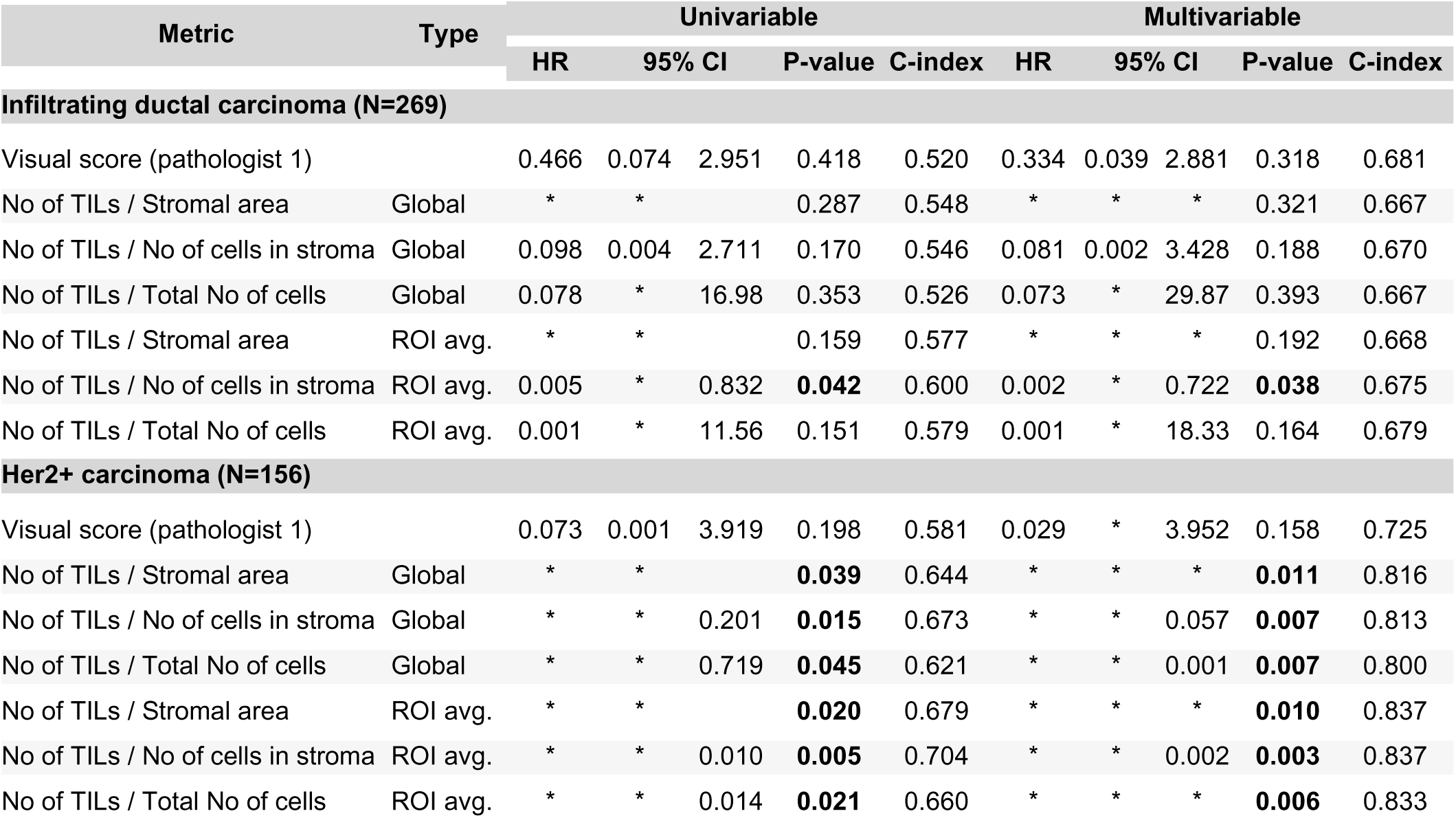
Cox regression survival analysis of the predictive value of visual and computational TILs scores for breast cancer progression. Each metric was combined with clinically salient covariates to create a separate multivariable model. All multivariable models were controlled for patient age and AJCC pathologic stage I and II status. Additionally, we controlled models using the infiltrating ductal carcinoma subset for basal genomic subtype status, and we controlled models using the Her2+ subset for infiltrating ductal histologic subtype status. Significant p-values are outlined in bold, using a significance threshold of 0.05. The * symbol indicates values < 0.001. Abbreviations used: HR, Hazard Ratio; 95%CI, upper and lower bounds of the 95% confidence interval; C-index, concordance index; No., number; Avg, weighted average.

We compared the performance of MuTILs to previously published models for tissue region segmentation [17] and nuclei instance segmentation [19]. The region segmentation performance of MuTILs was compared to the fully-convolutional network (VGG-FCN8) of [17] on common testing slides from both papers (see Supplement Table S8). We note that while MuTILs segments tissue regions at 10X objective magnification, the VGG-FCN8 model performs segmentation at 40X objective magnification. MuTILs improves segmentation of stromal regions while sacrificing some performance on epithelial and TIL regions (see Supplement Table S8). A per-slide performance comparison of tissue region segmentation is presented in Supplement Table S9. In nuclear classification, MuTILs performs better than the mask-RCNN model of [19] on all nuclei types, including by 2% for TIL nuclei (see Supplementary Table S9). As discussed earlier, for TILs scoring, the most clinically-relevant classes are stromal regions and TILs nuclei.

Computational TILs score variants had a modest to high correlation with the visual scores, with Spearman correlations ranging from 0.55 to 0.61 (all p-values < 0.001) (**Fig. 5**). Points in red are outliers that contributed to the correlation metric but were not used in calibration. Some slides were outliers with discrepant visual and computational scores; the causes for this discrepancy are discussed below. Both global and saliency-weighted scores were significantly correlated with the visual scores (p<0.001). We further analyzed pathologist-pathologist concordance using Bland-Altman analysis. For the pathologist-pathologist comparison, most all points fall within the +/− two standard deviations interval, with the strongest differences seen in the moderate scores ranging from 20%-60% with no evidence of proportional bias (see Supplement Figure S3). Score-score concordance was evaluated to measure agreement between scoring methods composed of score variants (nTSa, nTnS, nTnA) and score aggregation methods (global, saliency weighted). Correlations are high when comparing aggregation methods for the same score variant (Spearman, 0.89-0.92) and across score variants (0.72-0.86) (see Supplement Table S4).

**Fig 5.**
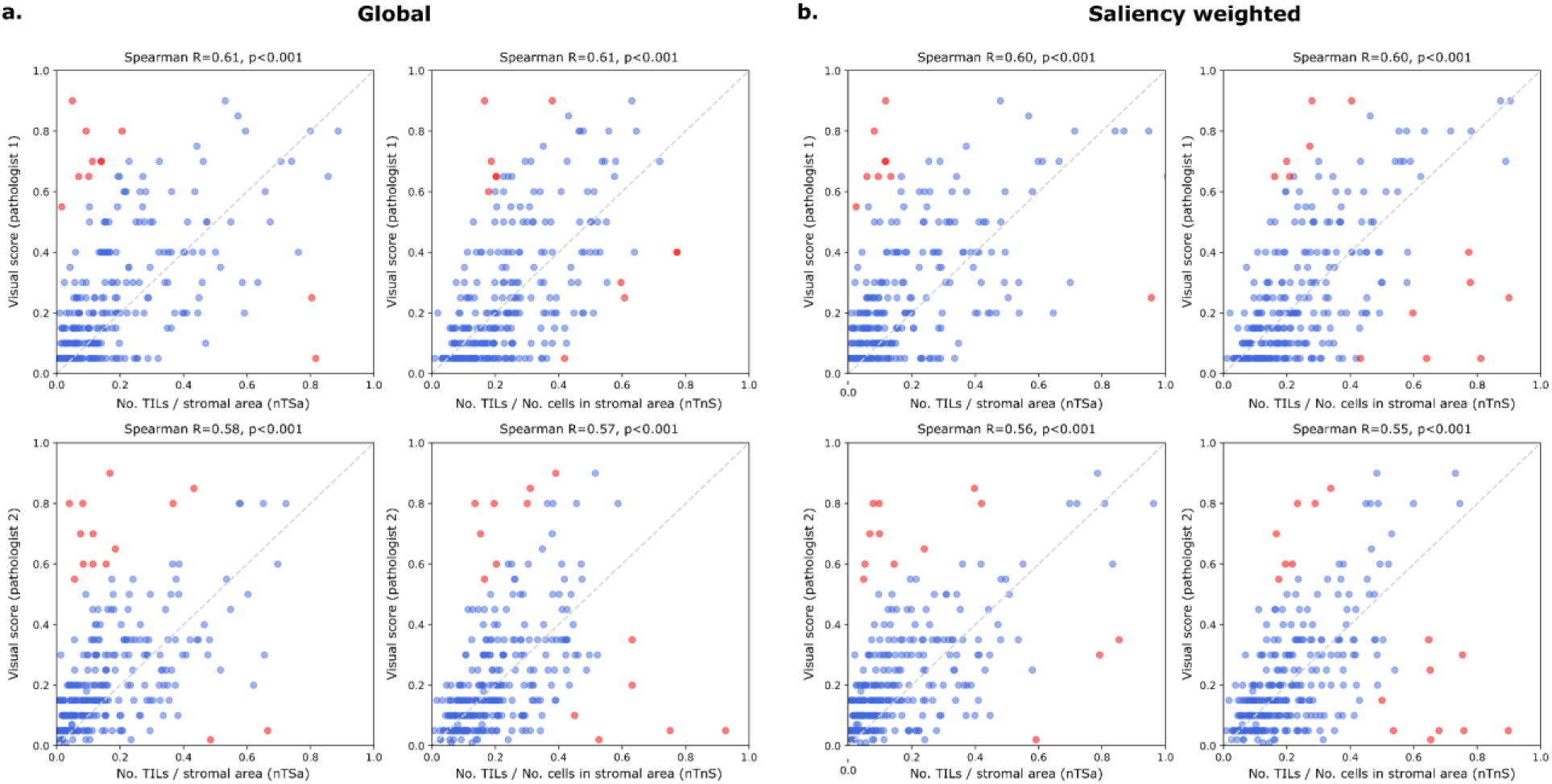
Correlation between visual and computational TILs scores. Visual scores were obtained from two pathologists using scoring recommendations from the TILs Working Group. Each point in the scatter plots above represents a single patient. Each plot above illustrates the correlation between the visual scores of one pathologist against either nTSa or nTnS scoring (using either global or salience weighted aggregation). Computational TIL scores were calibrated for the sake of interpretation to map them to a similar value range as the visual scores. Points in red are outliers that contributed to the correlation calculations but were not used during calibration. **a.** Scores obtained globally by aggregating data from all ROIs. **b.** Scores obtained by saliency-weighted averaging using estimated peritumoral stroma to weight each ROI.

We examined the prognostic value of MuTILs on infiltrating ductal carcinomas and Her2+ carcinomas (**Fig. 6**). While we had access to visual scores from the basal cohort, the number of outcomes was limited, and neither visual nor computational scores had prognostic value. All metrics were obtained by saliency-weighted averaging of computational scores from 300 ROIs. Both visual and computational scores had good separation within the infiltrating ductal cohort, although only the nTnS and nTnA computational scores had significant log-rank p-values (p=0.009 and p=0.006, respectively). Within the Her2+ cohort, all metrics had good separation on the Kaplan-Meier, although the visual score had a borderline p-value. All computational scores were significant within this cohort (p=0.018 for nTSa, p=0.002 for nTnS, and p=0.006 for nTnA).

**Fig 6.**
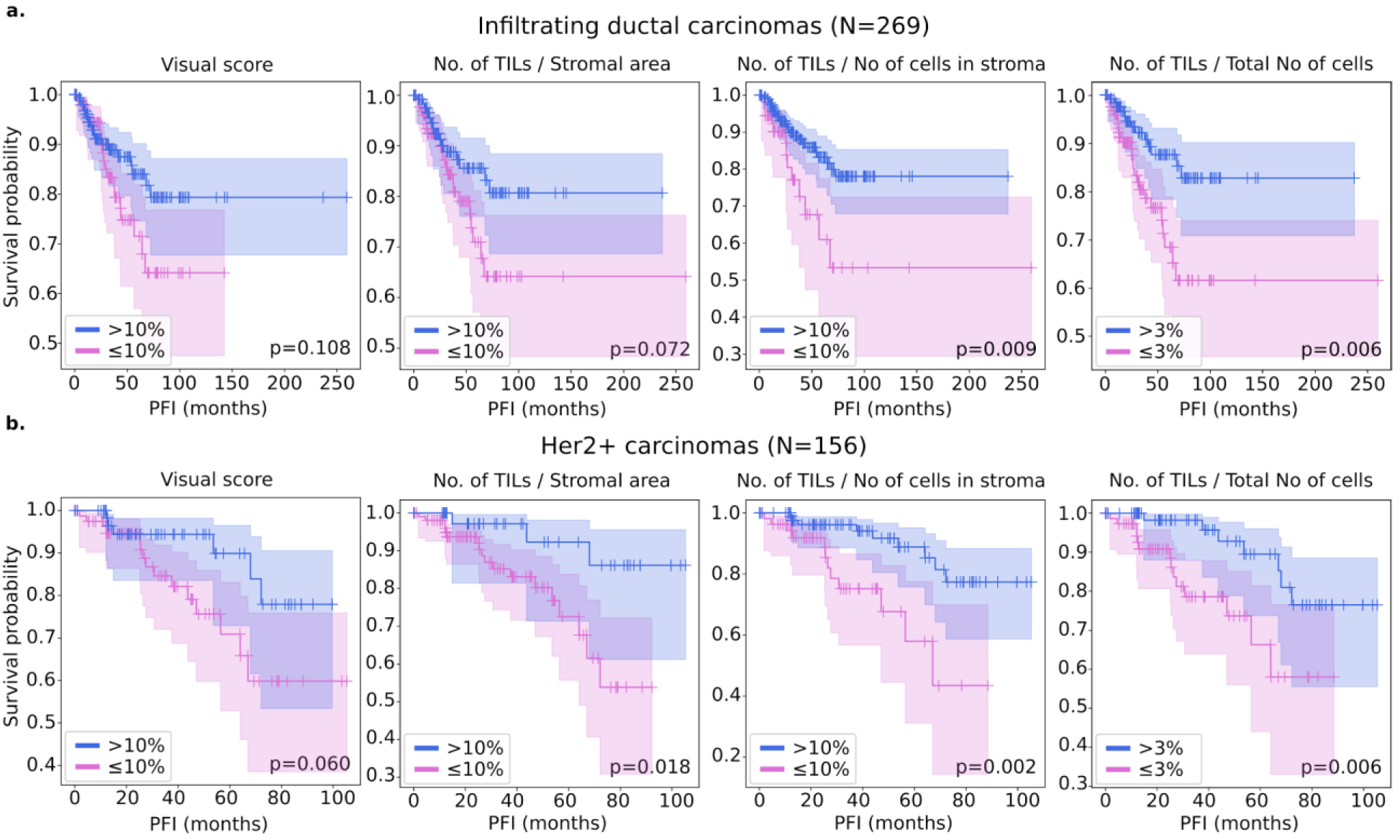
Kaplan-Meier analysis of visual and computational TILs assessment in predicting breast cancer progression. A threshold of 10% was used to define low-score and high-score patients for scores estimated in stromal regions which includes the visual, nTnS, and nTSa scores. For comparison, the nTnA score was included where the denominator includes all cells, not just those in the stromal compartment. For the nTnA score used a 3% threshold to account for the larger denominator. An analysis of the nTnA score distributions is presented in Figure S3 to justify this choice. We utilized visual scores from pathologist 1 as they are a breast cancer subspecialist. Both visual and computational scores effectively stratify outcomes in the infiltrating ductal and HER2+ carcinomas. Stratification for visual scores is clear but not statistically significant at the p=0.05 level. Computational scores generally improve stratification over visual scores and are statistically significant, except for the nTSa in infiltrating ductal carcinoma.

We also examined the prognostic value of the continuous (non-thresholded) TILs scores using Cox proportional hazards regression, with and without controlling for clinically-salient covariates, including patient age, AJCC pathologic stage, histologic subtype, and basal status (**Table 2**). The analysis was restricted to slides where visual TILs scores were available for a fair comparison. In the multivariable setting, a model was built for each metric combined with clinically salient covariates. We controlled all multivariable models for patient age and AJCC pathologic stage I and II status. Additionally, we controlled models using the infiltrating ductal carcinoma subset for basal genomic subtype status, and we controlled models using the Her2+ subset for infiltrating ductal histologic subtype status. Within the infiltrating ductal cohort, the only metric with significant independent prognostic value on multivariable analysis was the nTnS computational score. Within the Her2+ cohort, the visual score was not independently prognostic (p=0.158), while the computational scores all had independent prognostic value, with the most prognostic being the nTnS variant (p=0.003, HR<0.001). Saliency-weighted ROI scores almost always had better prognostic value than global computational scores.

## Discussion

One of the difficulties facing widespread adoption of state-of-the-art DL in medical domains is their opacity. There is a broad consensus that explainability is critical to trustworthiness, especially in clinical applications [1,12,27–29]. The standard application of DL models in histopathology involves the direct prediction of targets from the raw images. For example, we may predict patient survival given a WSI scan [30]. However, an alternative paradigm is beginning to emerge that combines the strong predictive power of opaque DL models and the interpretable nature of handcrafted features, a technique called Concept bottleneck modeling [31]. The fundamental idea is simple: 1. Use DL to delineate various tissue compartments and cells; 2. Extract handcrafted features that make sense to a pathologist; 3. Learn to predict the target variable, say patient survival, using an interpretable ML model that takes handcrafted features as its input. Hence, the most challenging task is handled using powerful DL models, while the terminal prediction task uses highly interpretable models.

MuTILs is a concept bottleneck model; it learns to predict the individual components that contribute to the TILs score (i.e., peritumoral stroma and TILs cells) and uses those to make the final predictions [31]. This setup makes its predictions explainable and helps identify sources of error. The region constraint helped provide context for the nuclear predictions at high resolution, which helped reduce the misclassification of immature fibroblasts and plasma cells as cancer (**Fig. 7**). To improve the reliability of tissue and cell classifications, we grouped model predictions into a simplified set of labels necessary for the core task of TIL quantification. For example, normal breast acini are not well represented in the training data, and so MuTILs model predictions are not reliable for distinguishing normal and cancer acini (Fig. 7, bottom row). Hence, we assessed performance at the level of grouped classes with reliable ground truth (epithelium, stroma, TILs) at evaluation. A richer set of predicted labels can be achieved by expanding the training set or downstream modeling of architectural patterns, which is beyond the scope of this work. The MuTILs model is able to analyze a typical slide in approximately 15 minutes on a server system equipped with multiple graphics processors. While this is competitive for a model that performs panoptic segmentation, additional speedup would need to be realized for clinical deployment.

**Fig 7.**
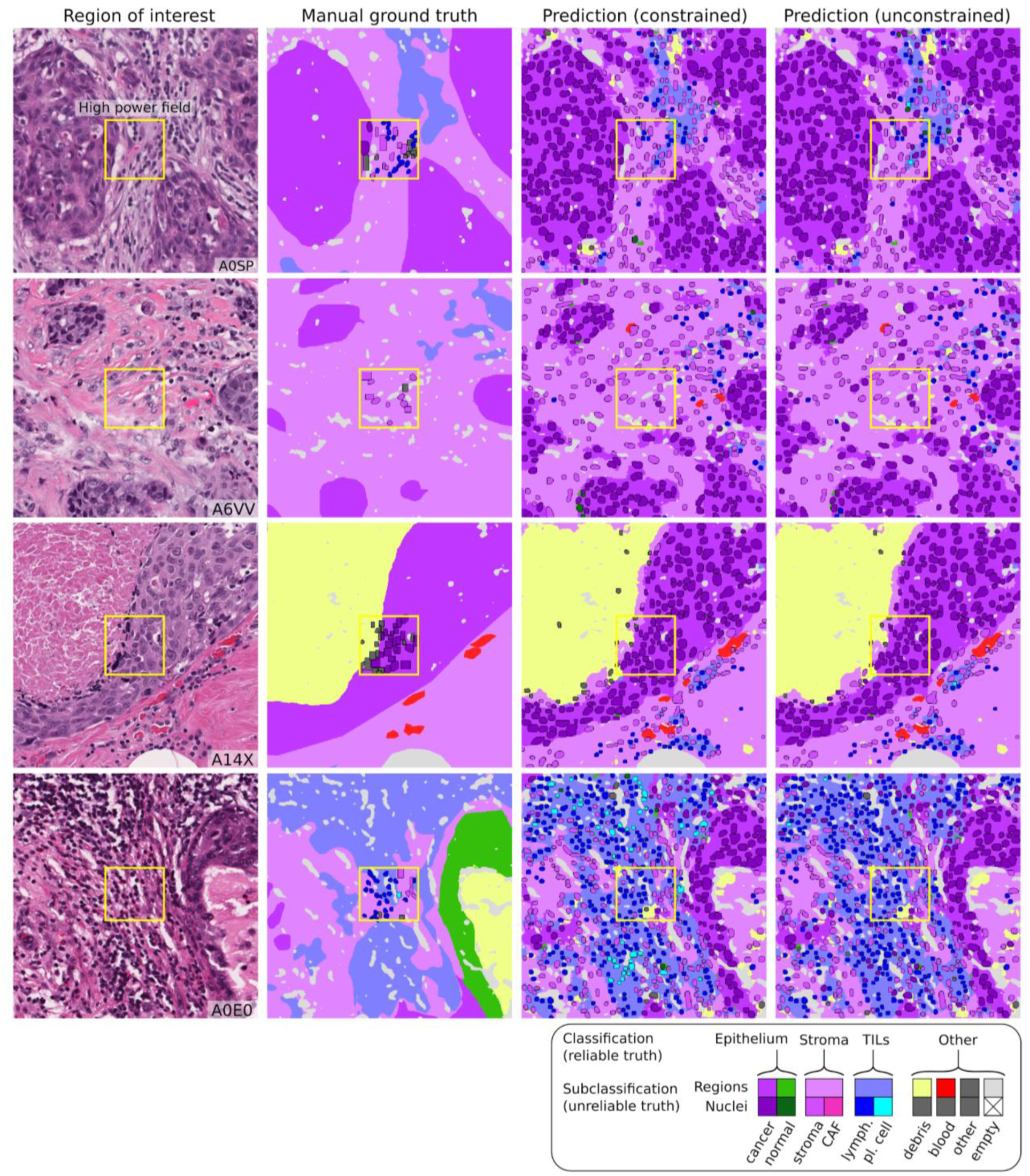
Qualitative examination of sample testing set predictions and sources of misclassification. The training set annotations of cells and tissue regions are very granular, for example distinguishing between lymphocytes and plasma cells. Some of these finer subclassifications have lower inter-rater agreement (“unreliable”), or are not abundant enough for a model to learn. Furthermore, the classes that a model needs to distinguish for the core task of TIL quantification is much simpler. Therefore, we assessed performance by grouping several classes to form a more reliable and practical ground truth (epithelium, stroma, TILs). The low abundance of normal breast acini in the training data makes it difficult for MuTILs models to distinguish normal and cancerous epithelial tissue (bottom row). We combine predictions of cancer and normal epithelium regions into a single “epithelium” class. Note how the region constraint improves nuclear classifications (third vs fourth column). This improvement is most notable for large TILs (first row) and immature fibroblasts (second row), which are misclassified as cancer without the region constraint.

A qualitative examination of slides with discrepant visual and computational TILs scores shows there are three major contributors to discrepancies:

1. Misclassifications of some benign or low-grade tumor nuclei as TILs.
2. Variations in TILs density in different areas within the slide, which cause inconsistencies in visual scoring. This phenomenon is also a well-known contributor to inter-observer variability in visual TILs scoring [11].
3. Variable influence of tertiary lymphoid structures on the WSI-level score.

Our results show that the most prognostic TILs score variant (nTnS) is derived from dividing the number of TILs cells by the total number of cells within the stromal region. The visual scoring guidelines rely on the nTSa, which is reflected in the slightly higher correlation of the nTSa variant with the visual scores compared to nTnS [10]. So why is nTnS more prognostic than nTSa? There are two potential explanations. First, it may be that nTnS is better controlled for stromal cellularity since it would be the same in low- vs. high-cellularity stromal regions as long as the proportion of stromal cells that are TILs is the same. Second, nTnS may be less noisy since it relies entirely on nuclear assessment at 20x objective, while stromal regions are segmented at half that resolution.

Finally, we note that this validation was done only using the TCGA cohort, and future work will include validation on more breast cancer cohorts. In addition, we note that MuTILs cannot distinguish cancer from normal breast tissue at low resolution, which may necessitate manual curation of the analysis region, especially for low-grade cases.

## Conclusion

We present PanopTILs, a segmentation dataset that enables the joint segmentation of tissue regions and cell nuclei. This dataset enabled us to train a novel panoptic segmentation model, MuTILs, which is a lightweight deep-learning model for reliable assessment of TILs in breast carcinomas in accordance with clinical scoring recommendations. It jointly classifies tissue regions and cell nuclei at different resolutions and uses these predictions to derive patient-level scores. We show that MuTILs can produce predictions with good generalization for the predominant tissue and cell classes relevant for TILs scoring. Furthermore, computational scores correlate significantly with visual assessment and have strong independent prognostic value in infiltrating ductal carcinoma and Her2+ cancer.

## Data Availability Statement

The PanopTILs dataset is made public at: https://sites.google.com/view/panoptils/.

## Code availability

Relevant code is publicly available at: github.com/PathologyDataScience/MuTILs_Panoptic

## Ethics statement

All data was shared with investigators in a deidentified form. All patients participated voluntarily and provided written informed consent. CPS-II data sharing was approved through the Emory University Institutional Review Board, approval number IRB00045780.

## Data Availability

The PanopTILs dataset is made public at: https://sites.google.com/view/panoptils/.
The BCSS and NuCLS datasets used for constructing PanopTILs are publicly available, and so are the TCGA clinical data. The Cancer Prevention Study-II data is available via the American Cancer Society (https://www.cancer.org/).

https://sites.google.com/view/nucls/home

https://github.com/PathologyDataScience/BCSS

## Acknowledgments

This work was supported by the U.S. NIH NCI grants U01CA220401 and U24CA19436201. We acknowledge support from Dr. David Gutman and the American Cancer Society, including Dr. Mia M. Gaudet, Dr. Samantha Puvanesarajah, Dr. Lauren Teras, James Hodge, and Elizabeth Bain.

## Author Contributions

**MA**: Idea conception, model implementation, validation, and manuscript review. **SL**: validation, manuscript writing. **MAR**: validation, manuscript writing. **RS**: manual scoring of TILs, manuscript approval. **LADC**: Idea conception, manuscript writing.

## Supplementary Material

**Supplementary Table S1.** Overview of nuclear annotations in PanopTILs dataset.

| Nuclei annotation type | Count |
| --- | --- |
| Cancer | 16322 |
| Lymphocyte | 9596 |
| Fibroblast | 6945 |
| Debris | 5943 |
| Plasma Cell | 4641 |
| Active Stromal Cell | 1041 |
| Normal epithelium | 382 |
| Other Cell | 3 |

**Supplementary Table S2.** Impact of region constraint on accuracy of nuclei classifications. Bolded values indicate higher performance. Utilizing region constraints improves accuracy for most categories in most test folds. Notably, region constraints improve classification accuracy for the Epithelium and Fibroblast classes.

|  | Fold1 | Fold2 | Fold3 | Fold4 | Fold5 |
| --- | --- | --- | --- | --- | --- |
| **Accuracy with region constraint** |  |  |  |  |  |
| Epithelial | **87.3** | **84.2** | **92.5** | **88.1** | **88.4** |
| Stromal | **84.7** | **81.1** | **87.2** | **82.4** | **84.0** |
| TIL | **90.7** | **89.9** | **92.0** | **88.9** | **90.8** |
| Debris | **95.5** | 93.9 | **97.1** | **95.5** | 97.0 |
| **Accuracy without region constraint** |  |  |  |  |  |
| Epithelial | 83.2 | 83.6 | 91.6 | 83.2 | 83.7 |
| Stromal | 82.7 | 81.0 | 87.1 | 79.0 | 82.5 |
| TIL | 89.5 | 88.6 | 91.8 | 86.9 | 90.0 |
| Debris | 95.4 | **95.8** | 96.8 | 95.2 | **97.4** |

**Supplementary Table S3.** Impact of region constraint on Matthews Correlation Coefficient of nuclei classifications. Bolded values indicate higher performance. Utilizing region constraints improves MCC for all categories in all test folds. Using region constraints improves MCC for Epithelial and Fibroblast classes. Performance improvements for debris are considerable.

|  | Fold1 | Fold2 | Fold3 | Fold4 | Fold5 |
| --- | --- | --- | --- | --- | --- |
| **MCC with region constraint** |  |  |  |  |  |
| **Epithelial** | **74.4** | **69.5** | **84.6** | **73.5** | **76.3** |
| **Stromal** | **60.5** | **54.4** | **67.4** | **57.7** | **61.0** |
| **TIL** | **74.9** | **73.7** | **80.0** | **75.7** | **77.0** |
| **Debris** | **36.1** | **35.5** | **57.2** | **44.4** | **27.7** |
| **MCC without region constraint** |  |  |  |  |  |
| **Epithelial** | 66.9 | 67.2 | 82.9 | 64.2 | 67.9 |
| **Stromal** | 53.4 | 53.7 | 67.0 | 48.3 | 55.2 |
| **TIL** | 72.3 | 71.4 | 79.7 | 72.0 | 75.0 |
| **Debris** | 17.5 | 0.0 | 44.6 | 1.7 | -0.2 |

**Supplementary Table S4.** Impact of region constraint on AUROC of nuclei classifications. Bolded values indicate higher performance. Utilizing region constraints improves MCC for all categories in all test folds.

|  | Fold1 | Fold2 | Fold3 | Fold4 | Fold5 |
| --- | --- | --- | --- | --- | --- |
| **AUROC with region constraint** |  |  |  |  |  |
| **Epithelial** | **94.0** | **93.7** | **97.3** | **94.0** | **95.5** |
| **Stromal** | **90.0** | **87.7** | **93.2** | **88.3** | **90.4** |
| **TIL** | **96.2** | **96.4** | **97.6** | **96.2** | **96.8** |
| **Debris** | **84.6** | **86.2** | **93.4** | **89.9** | **87.7** |
| **AUROC without region constraint** |  |  |  |  |  |
| **Epithelial** | 91.0 | 91.6 | 97.0 | 90.4 | 92.6 |
| **Stromal** | 87.9 | 85.9 | 92.9 | 84.4 | 88.4 |
| **TIL** | 95.3 | 95.5 | 97.5 | 94.8 | 96.0 |
| **Debris** | 80.0 | 75.9 | 93.3 | 73.8 | 77.4 |

**Supplementary Table S5.** Impact of region constraint on precision and recall of nuclei classifications. Results are shown in precision/recall pairs. Bolded values indicate higher performance (average of precision and recall). In many instances, using region constraints improves precision markedly with only a small tradeoff in recall.

|  | Fold1 | Fold2 | Fold3 | Fold4 | Fold5 |
| --- | --- | --- | --- | --- | --- |
| **Precision / recall with region constraint** |  |  |  |  |  |
| **Epithelial** | **84.4 / 87.7** | **92.9 / 71.6** | **90.8 / 91.5** | **84.4 / 80.5** | **84.1 / 88.2** |
| **Stromal** | **71.3 / 70.5** | **61.8 / 73.3** | **71.2 / 81.0** | **74.0 / 66.3** | **69.6 / 75.1** |
| **TIL** | **80.1 / 82.1** | **72.8 / 88.2** | **89.9 / 81.2** | **75.3 / 92.2** | **87.0 / 79.5** |
| **Debris** | **49.8 / 29.4** | **32.9 / 45.3** | **62.5 / 55.3** | **53.8 / 40.6** | **38.2 / 22.1** |
| **Precision / recall without region constraint** |  |  |  |  |  |
| **Epithelial** | 77.4 / 88.0 | 87.5 / 75.4 | 87.9 / 92.8 | 73.2 / 81.6 | 75.4 / 88.7 |
| **Stromal** | 70.7 / 59.0 | 61.7 / 72.1 | 71.4 / 80.0 | 70.9 / 54.5 | 70.2 / 64.2 |
| **TIL** | 76.0 / 82.7 | 69.0 / 89.3 | 90.2 / 80.4 | 71.3 / 91.9 | 84.2 / 79.6 |
| **Debris** | 44.8 / 8.0 | 0.0 / 0.0 | 64.1 / 33.0 | 29.2 / 0.1 | 0.0 / 0.0 |

**Supplementary Table S6.** Impact of region constraint on F1 score of nuclei classifications. Bolded values indicate higher performance. Utilizing region constraints improves F1 score for all categories in all test folds except for 1 (Fold 2, Epithelial), where the difference is only 0.01.

|  | Fold1 | Fold2 | Fold3 | Fold4 | Fold5 |
| --- | --- | --- | --- | --- | --- |
| **F1 score with region constraint** |  |  |  |  |  |
| **Epithelial** | **86.0** | 80.9 | **91.1** | **82.4** | **86.1** |
| **Stromal** | **70.9** | **67.1** | **75.8** | **69.9** | **72.2** |
| **TIL** | **81.1** | **79.8** | **85.3** | **82.9** | **83.1** |
| **Debris** | **37.0** | **38.1** | **58.6** | **46.3** | **28.0** |
| **F1 score without region constraint** |  |  |  |  |  |
| **Epithelial** | 82.4 | **81.0** | 90.3 | 77.2 | 81.5 |
| **Stromal** | 64.3 | 66.5 | 75.4 | 61.6 | 67.0 |
| **TIL** | 79.2 | 77.8 | 85.0 | 80.3 | 81.9 |
| **Debris** | 13.6 | 0.0 | 43.6 | 0.3 | 0.0 |

**Supplementary Table S7.** Impact of region constraint on specificity and sensitivity of nuclei classifications. Bolded values indicate higher performance (average of sensitivity and specificity). Utilizing region constraints often improves sensitivity with a modest tradeoff in specificity for most classes and in most folds.

|  | Fold1 | Fold2 | Fold3 | Fold4 | Fold5 |
| --- | --- | --- | --- | --- | --- |
| **Specificity / sensitivity with region constraint** |  |  |  |  |  |
| Epithelial | **86.9 / 87.7** | **95.2 / 71.6** | **93.2 / 91.5** | **92.1 / 80.5** | **88.6 / 88.2** |
| Stromal | **89.8 / 70.5** | **83.9 / 73.3** | **89.3 / 81.0** | **89.6 / 66.3** | **87.4 / 75.1** |
| TIL | **93.5 / 82.1** | **90.4 / 88.2** | **96.3 / 81.2** | **87.6 / 92.2** | **95.3 / 79.5** |
| Debris | **98.6 / 29.4** | **96.0 / 45.3** | **98.7 / 55.3** | **98.2 / 40.6** | **99.0 / 22.1** |
| **Specificity / sensitivity without region constraint** |  |  |  |  |  |
| Epithelial | 79.3 / 88.0 | 90.6 / 75.4 | 90.6 / 92.8 | 84.1 / 81.6 | 80.3 / 88.7 |
| Stromal | 91.2 / 59.0 | 84.1 / 72.1 | 89.5 / 80.0 | 90.0 / 54.5 | 89.5 / 64.2 |
| TIL | 91.7 / 82.7 | 88.4 / 89.3 | 96.5 / 80.4 | 84.8 / 91.9 | 94.1 / 79.6 |
| Debris | 99.5 / 8.0 | 100 / 0.0 | 99.3 / 33.0 | 100 / 0.1 | 100 / 0.0 |

**Supplementary Table S8.** Performance comparison of tissue region segmentation of MuTILs versus the VGG-FCN8 model described in [17]. Bolded values indicate higher performance. For a fair comparison only slides present in the testing set(s) of both models were used. Note that VGG-FCN8 model segments slides at a 40x magnification, while MuTILs is trained to segment slides at a 10x magnification to provide low-power context for the nucleus classifications. While this results in some drop in accuracy of the cancer and TILs-dense region segmentation, the lower power context actually improves stromal region classification.

|  | Epithelial | Stromal | TILs |
| --- | --- | --- | --- |
| **MuTILs tissue segmentation (10X magnification)** |  |  |  |
| DICE overall | 86.8 | **85.9** | 70.2 |
| DICE slide average (std) | 82.1 (13.2) | **82.3 (8.9)** | 62.4 (21.4) |
| **VGG-FCN8 tissue segmentation (40X magnification)** |  |  |  |
| DICE overall | **89.1** | 82.2 | **77.3** |
| DICE slide average (std) | **86.8 (7.9)** | 77.9 (12.2) | **70.0 (22.6)** |

**Supplementary Table S9.** Performance comparison of nuclei classification of MuTILs versus the mask-RCNN model described in [19]. Bolded values indicate higher performance. Assignment to training/testing folds was the same in both works, allowing exact comparison. Mean and standard deviation statistics exclude fold 1, which contributed to model turning. MuTILs outperforms the mask-RCNN model for all nuclei types evaluated.

|  | Fold1 | Fold2 | Fold3 | Fold4 | Fold5 | Mean (std) |
| --- | --- | --- | --- | --- | --- | --- |
| **MuTILs MCC** |  |  |  |  |  |  |
| **Epithelial** | **74.4** | 69.5 | **84.6** | 73.5 | **76.3** | **76.0 (6.4)** |
| **Stromal** | **60.5** | **54.4** | **67.4** | **57.7** | **61.0** | **60.1 (5.5)** |
| **TIL** | **74.9** | 73.7 | **80.0** | 75.7 | **77.0** | **76.6 (2.6)** |
| **mask-RCNN MCC** |  |  |  |  |  |  |
| **Epithelial** | 72.9 | **73.7** | 74.9 | **80.6** | 57.4 | 71.7 (10.0) |
| **Stromal** | 47.1 | 53.0 | 46.9 | 56.9 | 40.7 | 49.4 (7.1) |
| **TIL** | 73.7 | **76.6** | 77.9 | **79.6** | 60.1 | 73.5 (9.1) |
| **MuTILs AUROC** |  |  |  |  |  |  |
| **Epithelial** | 94.0 | 93.7 | **97.3** | 94.0 | **95.5** | **95.1 (1.7)** |
| **Stromal** | **90.0** | **87.7** | **93.2** | 88.3 | **90.4** | **89.9 (2.5)** |
| **TIL** | **96.2** | **96.4** | **97.6** | **96.2** | **96.8** | **96.8 (0.6)** |
| **mask-RCNN AUROC** |  |  |  |  |  |  |
| **Epithelial** | **94.2** | **94.5** | 96.1 | **97.2** | 88.8 | 94.2 (3.7) |
| **Stromal** | 83.2 | 87.4 | 84.3 | **89.1** | 80.7 | 85.4 (3.7) |
| **TIL** | 95.3 | 96.2 | 95.7 | 95.9 | 91.0 | 94.7 (2.4) |

**Supplementary Figure S1.**
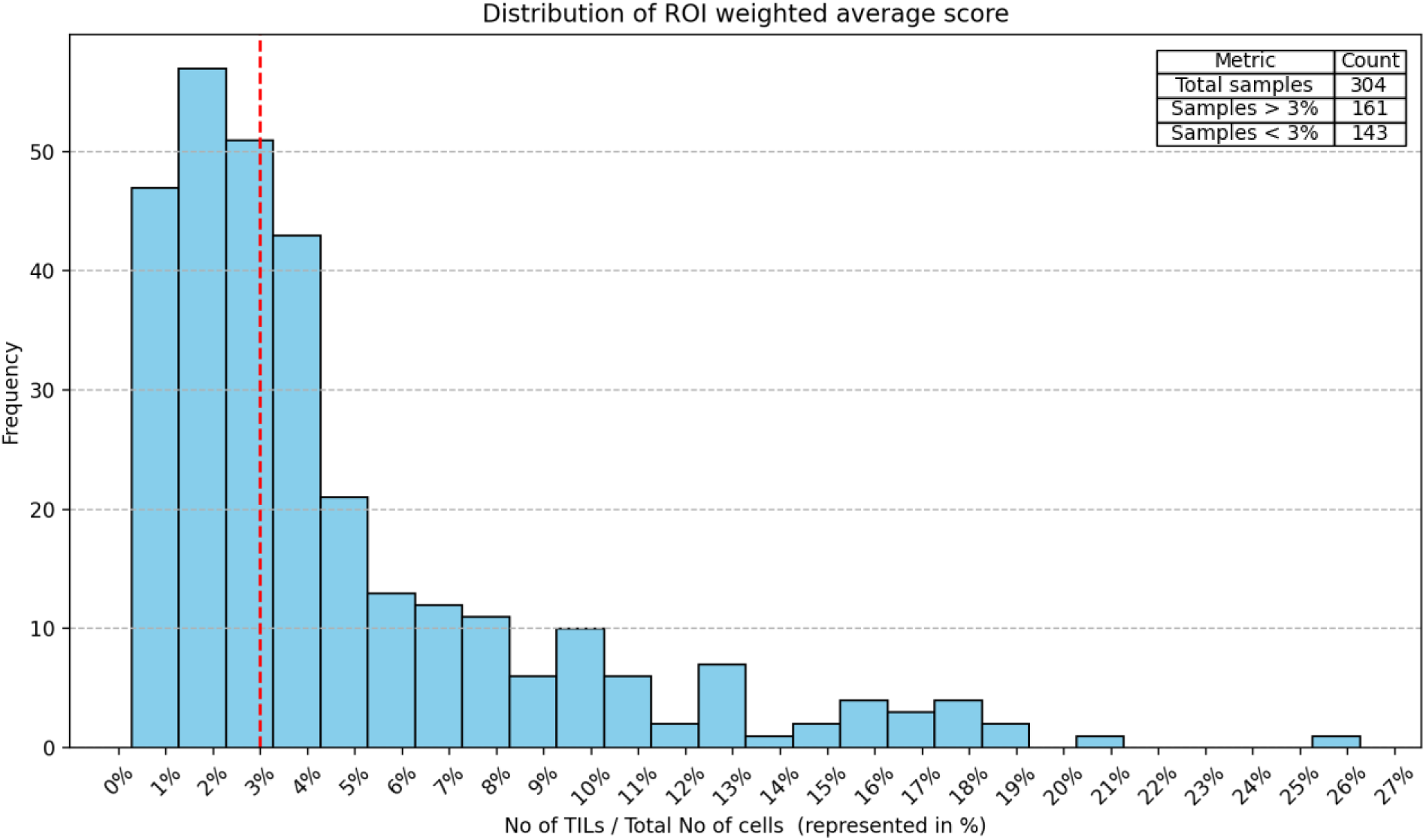
Distribution of No of TILs / Total No of cells. The distribution of the “No of TILs / Total No of cells” (nTnA) TIL Score. This score includes all cells in the denominator, and so the 10% threshold used for stromal scores is too conservative. The three leftmost histogram bins encompass around 50% of the total patients. Based on our observation, we selected a threshold value of 3% as it roughly represents the midpoint of this cumulative distribution where half of the patients lie. A 10% threshold would result in a significant imbalance and make comparison between the different scores difficult. The accompanying summary table further emphasizes the distribution of samples, highlighting that out of 304 total samples, 161 samples are above the 3% mark and 143 samples are below it.

**Supplementary Figure S2.**
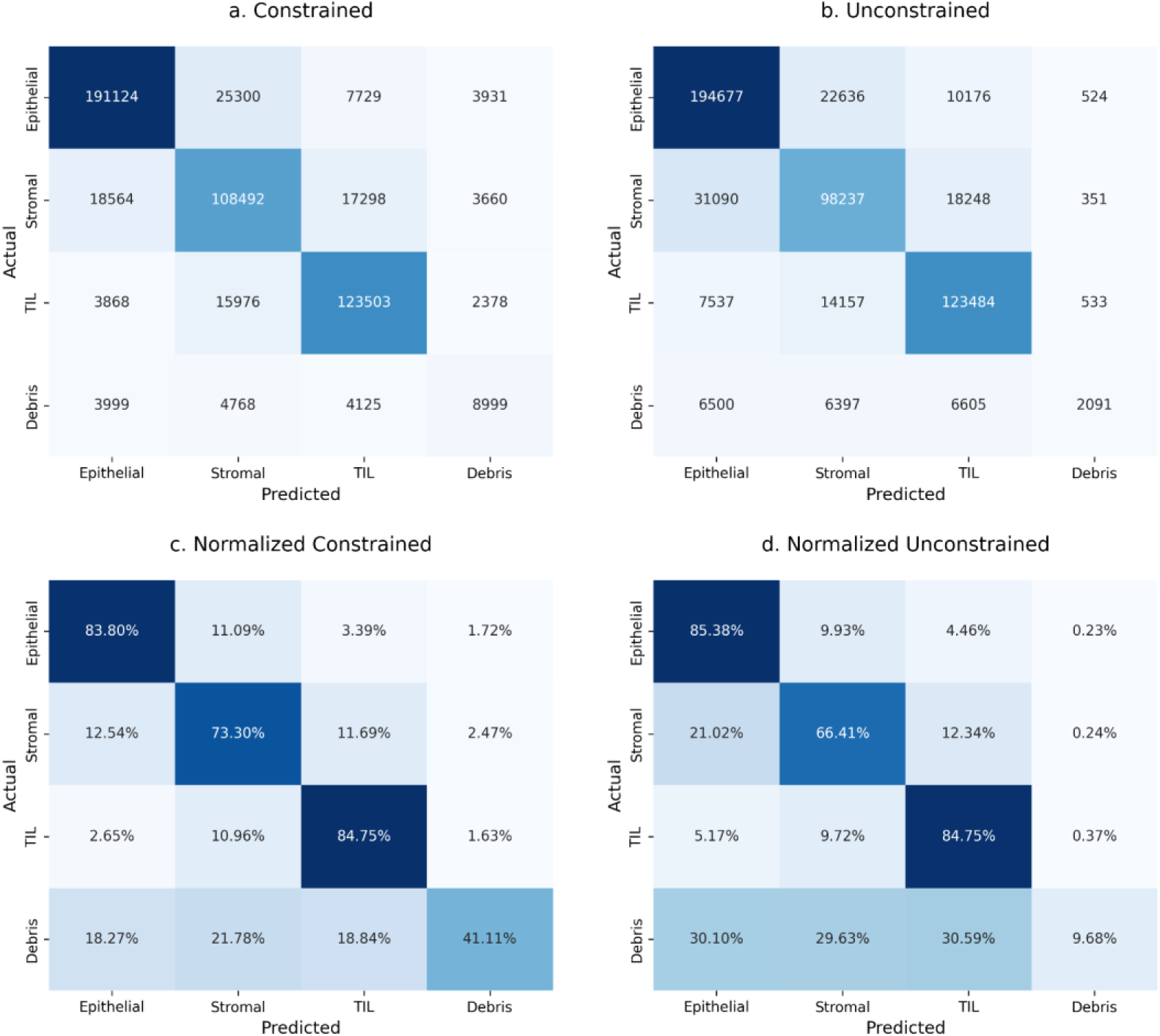
Confusion matrix of MuTIL nuclear classification. Values represent predictions aggregated over all samples and validation folds. Plots in the top row present classification counts where plots in the bottom row present percentages calculated for each ground truth label. **a.** The region constraint improves classification of stromal (fibroblast) and debris nuclei. **b.** Without region constraint, classification of epithelial nuclei improves at the cost of misclassifications for stromal and debris nuclei.

**Supplementary Figure S3.**
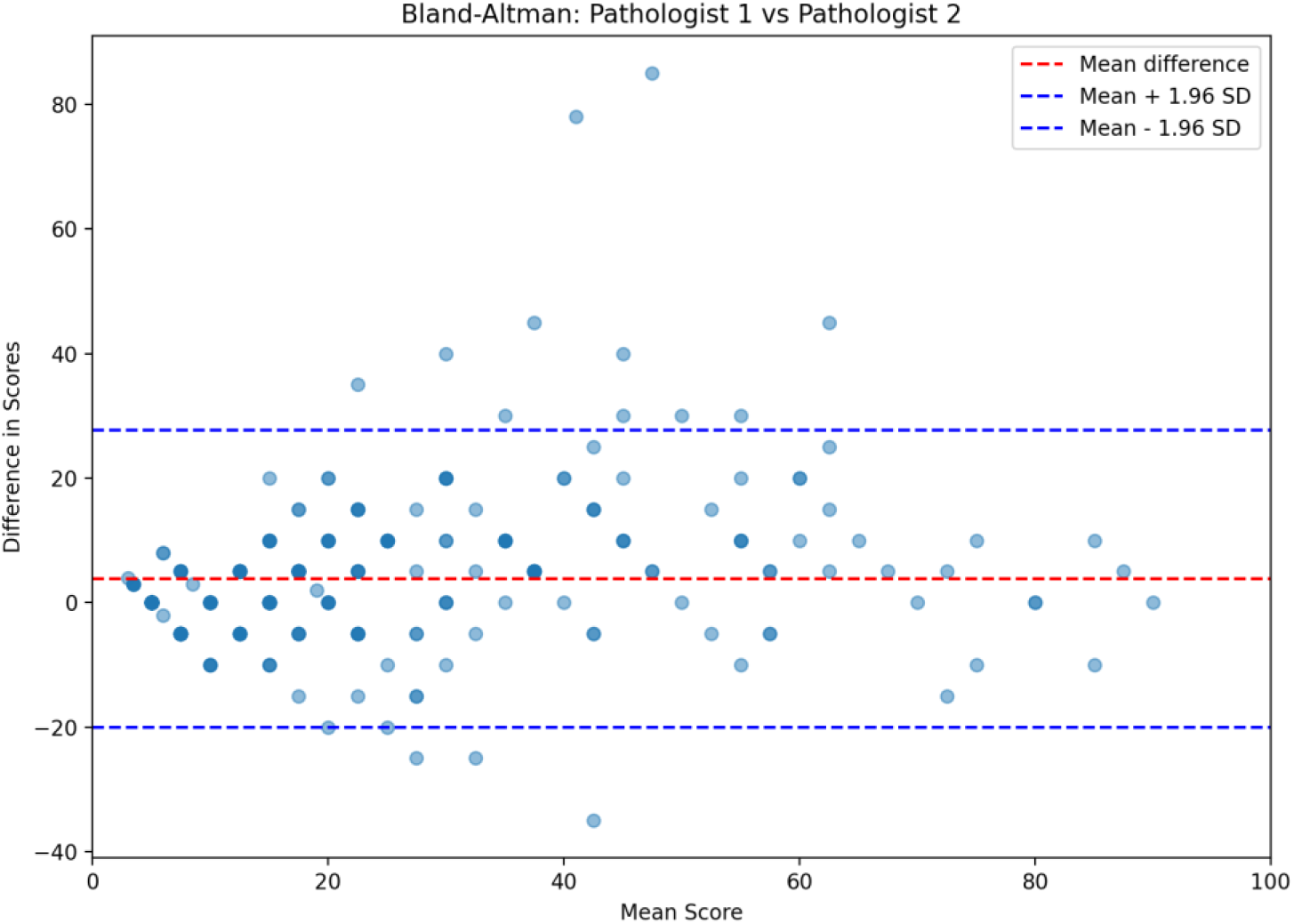
Bland-Altman plots between pathologists. Most points lie within +/− two standard deviation interval. Outliers are in the moderate score range from %20-%60. Most outliers indicate a higher bias for pathologist 2. No proportional bias is observed in higher scoring cases.

**Supplementary Figure S4.**
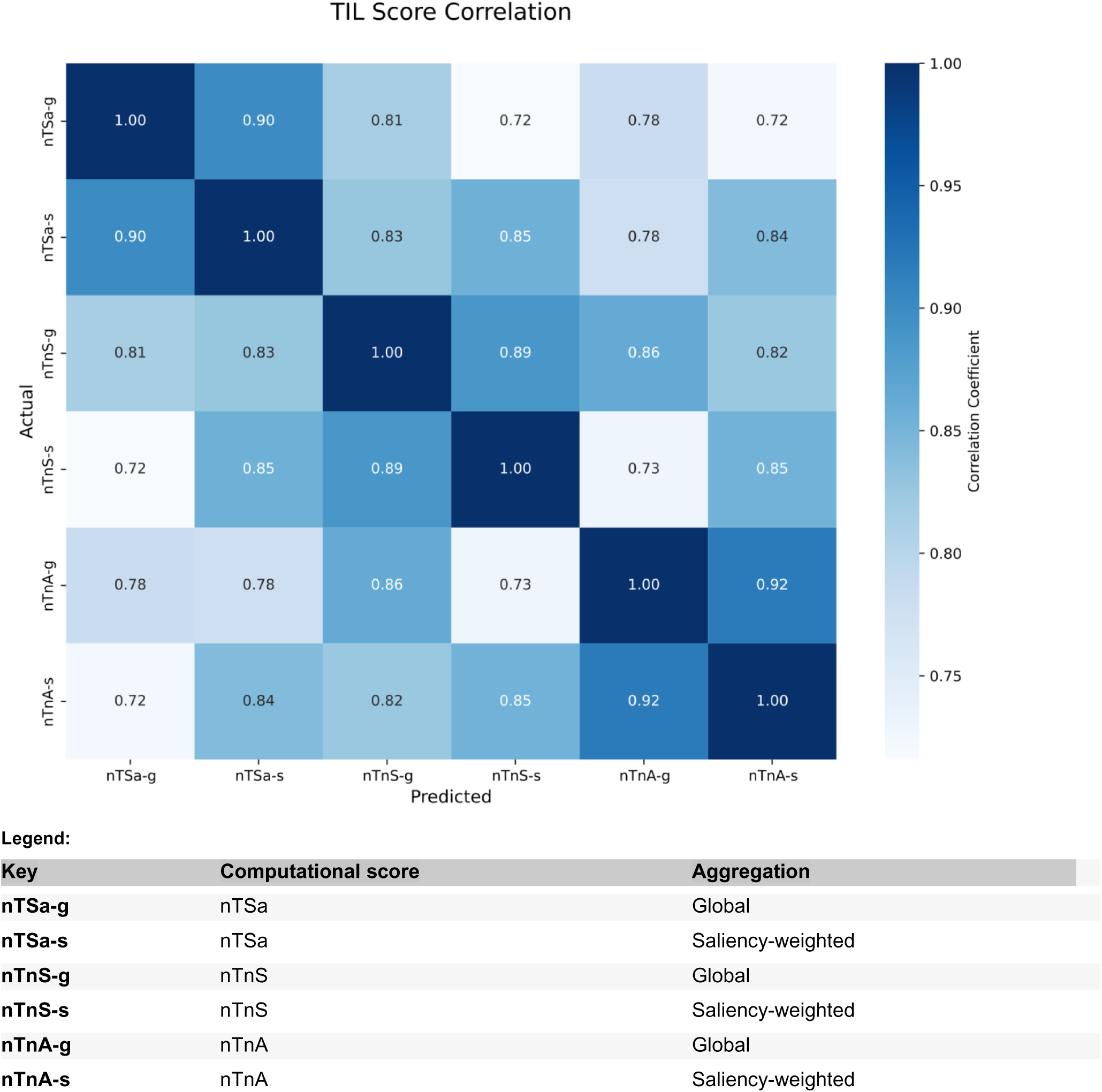
Heatmap computational TIL score variant correlation. Spearman correlations were calculated for each combination of score variants (nTSa, nTnS, nTnA) and score aggregation method (global, saliency weighted). For each variant, correlations between the global and saliency weighted scores are high, ranging from 0.89-0.92. Across variants, correlations are lower but still high, ranging from 0.72 to 0.86. It’s noteworthy that within each scoring category—whether focusing on stromal area, number of cells in stroma, or total number of cells—the global and ROI average scores consistently show high correlation. This highlights the reliability and coherence of the TIL score measurements.

